# Analysis of SARS-CoV-2 Ig seroprevalence in Northern Ireland

**DOI:** 10.1101/2023.09.19.23295776

**Authors:** Michelle K Greene, Peter Smyth, Andrew English, Joseph McLaughlin, Magda Bucholc, Janice Bailie, Julie McCarroll, Margaret McDonnell, Alison Watt, George Barnes, Mark Lynch, Kevan Duffin, Gerard Duffy, Claire Lewis, Jacqueline A James, Tom Ford, Maurice O’Kane, Taranjit Singh Rai, Anthony J Bjourson, Christopher Cardwell, J Stuart Elborn, David S Gibson, Christopher J Scott

## Abstract

**Background:** With the impact of SARS-CoV-2 upon public health directly and socioeconomically, further information was required to inform policy decisions designed to limit virus spread. This study sought to contribute to serosurveillance work within Northern Ireland to track SARS-CoV-2 progression and guide health strategy.

**Methods:** Sera/plasma samples from clinical biochemistry laboratories were analysed for anti-SARS-CoV-2 immunoglobulins (Ig). Samples were assessed using an Elecsys anti-SARS-CoV-2 or anti-SARS-CoV-2 S ECLIA (Roche) on an automated Cobas-e-analyser. Samples were also assessed via ELISA (Euroimmun). A subset of samples assessed via Roche Elecsys anti-SARS-CoV-2 IgG assay were subsequently analysed in an ACE2 pseudoneutralisation assay using a V-PLEX SARS-CoV-2 Panel 7 for IgG and ACE2 by MesoScale Diagnostics Inc.

**Results:** Across three testing rounds (June-July 2020, November-December 2020 and June-July 2021 (rounds 1-3 respectively)), 4844 residual sera/plasma specimens were assayed for SARS-CoV-2 Ig. Seropositivity rates increased across the study, peaking at 11.6% during round 3. Varying trends in SARS-CoV-2 seropositivity were noted based on demographic factors. For instance, highest rates of seropositivity shifted from older to younger demographics across the study period. In round 3, alpha (B.1.1.7) variant neutralising antibodies were most frequently detected across age groups, with median concentration of anti-spike protein antibodies elevated in 50-69 year olds and anti-S1 RBD antibodies elevated in over 70s, relative to other age groups.

**Conclusions:** With seropositivity rates of <15% across the assessment period, it can be concluded that the significant proportion of the Northern Ireland population had not yet naturally contracted the virus by mid-2021.

## Introduction

Since the first reports of SARS-CoV-2, countless individuals have contracted the disease worldwide leading to significant mortality, morbidity and socioeconomic implications. Efforts to control SARS-CoV-2 transmission led the UK government to impose nationwide lockdowns, strict social distancing measures and widescale testing programmes to rapidly identify and isolate infected individuals. Despite these restrictions, continued spread of SARS-CoV-2 remained a challenge, highlighting the need for ongoing surveillance to track disease epidemiology and guide public health policy moving forwards.

Numerous options exist for testing and monitoring of SARS-CoV-2, including the reverse transcription polymerase chain reaction (RT-PCR) that has formed the gold standard diagnostic approach to date.^1–3^ This highly sensitive technique facilitates the molecular detection of viral RNA that is typically collected via nasopharyngeal swabs, although it is prohibited by the need for significant laboratory infrastructure and longer turnaround times. Moreover, RT-PCR is of limited use beyond the acute phase of SARS-CoV-2 infection so prevalence rates may be grossly underestimated. Alternatively, serological assays enable the detection of SARS-CoV-2-reactive antibodies in sera/plasma, which are released into the circulation in the weeks following infection and often persist for many months thereafter.^4–6^ These assays therefore provide an indication of prior exposure to SARS-CoV-2 and are particularly insightful for gathering information on asymptomatic infection in those who may not have satisfied eligibility criteria for PCR screening, providing a more refined estimate of SARS-CoV-2 prevalence. Many serological studies have been conducted worldwide^7–12^ and have informed planning in several contexts (e.g. vaccination scheduling in response to waning immunity), highlighting the pivotal contribution of serosurveillance towards control and containment of SARS-CoV-2.

For Northern Ireland in particular, estimates of SARS-CoV-2 seroprevalence have been largely derived from the ongoing Coronavirus (COVID-19) Infection Survey, where sera are sampled from randomly selected households across each of the jurisdictions within the UK. Here, we employed an alternative sampling source to derive a further independent estimate of the proportion of Northern Irish residents who contracted SARS-CoV-2 in the period to July 2021. The Northern Ireland health system is divided into five Health and Social Care Trusts and therefore to efficiently sample a broad spectrum of the population, residual sera/plasma specimens were procured from clinical biochemistry laboratories within each of the five Trusts and analysed for the presence of anti-SARS-CoV-2 immunoglobulins (Ig) using commercial immunoassay kits in both hospital and academic settings. We report temporal and demographic trends in Ig response, providing crucial insight into how seropositivity has evolved over time and how it has been influenced by characteristics including age, gender and place of residence.

## Methods

### Sample collection

Ethical approval for this study was obtained from the Northern Ireland Biobank. The Northern Ireland Biobank has ethical approval from The Office of Research Ethics Committees Northern Ireland (ORECNI REF 21/NI/0019) and can confer ethical approval for projects which have received material from the bank.^13^ Residual blood sera/plasma specimens (originally collected and processed for other clinical purposes) were sourced from Clinical Biochemistry laboratories within the various Health and Social Care Trusts across Northern Ireland, comprising the Belfast, Northern, Southern, South Eastern and Western Health and Social Care Trusts. Samples were acquired during three timeframes: June-July 2020, November-December 2020 and June-July 2021.

### Anti-SARS-CoV-2 IgG electrochemiluminescence immunoassay (ECLIA)

Sera/plasma samples were assayed using the Elecsys anti-SARS-CoV-2 or anti-SARS-CoV-2 S ECLIA (Roche) on an automated Cobas e analyser in accordance with the manufacturer’s instructions.

### Anti-SARS-CoV-2 IgG enzyme-linked immunosorbent assay (ELISA)

Sera/plasma samples were assayed using the anti-SARS-CoV-2 IgG ELISA (Euroimmun) in accordance with the manufacturer’s instructions. Semiquantitative assessment of anti-SARS-CoV-2 IgG was achieved by calculating the ratio of the extinction of the control or donor sample over the extinction of the calibrator. Ratios were interpreted as follows: < 0.8 = negative, ≥ 0.8 to < 1.1 = borderline and ≥ 1.1 = positive.

### ACE2 pseudoneutralisation IgG assay

A subset of n=278 plasma samples collected in the June-July 2021 cohort with Roche Elecsys anti-SARS-CoV-2 IgG assay data (n=219 S-antibody reactive samples; n=59 S-antibody negative samples as controls) were subsequently analysed by an ACE2 pseudoneutralisation assay. Receptor Binding Domain (RBD)-specific IgG and their neutralising capacity against SARS-CoV-2 wild type and variants B.1.1.7 (Alpha), B.1.351 (Beta) and P.1 (Gamma) were measured using the V-PLEX SARS-CoV-2 Panel 7 for IgG and ACE2 by MesoScale Diagnostics Inc. (MSD, USA) according to the manufacturer’s instructions and expressed as neutralising antibody concentration in μg/ml. Briefly, the assay uses spot multiplexing of 9 viral antigens within each plate well. For the ACE2 neutralisation assay, the ability of antibodies to block the binding of ACE2 to its cognate ligands was measured. Antibodies in plasma samples (diluted 1:10) that block the RBD antigen interaction with ACE2 on the spots were analysed, and a recombinant human ACE2 conjugated with MSD SULFO-TAG™ was used for detection. All plates were analysed on a MESO QuickPlex SQ 120 electrochemiluminescence reader to measure the light emitted from the SULFO-TAG™. Results for the MSD assays are reported in μg/ml derived from back fitting the measured signals for samples to a calibration curve generated for each plate. For the 7-point calibration curve, a 4-fold serial dilution was used with two replicates at each calibrator level and an additional zero calibrator blank. By correcting for dilution, the final antibody concentrations in undiluted samples were obtained. Detailed information on antigens, control samples and ACE2 calibrator reagent, including concentrations in μg/ml, are presented in the kit inserts provided by MSD (ACE2 cat no. K15440U). Cut off values supplied by the manufacturer of 0.762 μg/ml and 0.533 μg/ml were used to determine participant reactivity to SARS-CoV-2 S1 RBD and SARS-CoV-2 spike, respectively.

### Data analysis

Crude and standardised apparent proportions with anti-SARS-CoV-2 Ig were determined along with 95% confidence intervals. Additionally, crude and standardised true proportions were determined adopting the Bayesian method used by Public Health England^14^, which accounted for the sensitivity of 71/100 (71%) and specificity of 777/786 (98.9%) of the test. Crude true proportion was also determined using the Rogan-Gladen method^15^ to account for test sensitivity and specificity and gave similar results (analysis not shown). GraphPad Prism software (version 9.1.1) was used to plot data. Variables including age, gender, Health and Social Care Trust and postcode area were analysed using descriptive statistics. Frequencies and percentages were calculated for categorical variables and median and inter-quartile range for continuous variables. The chi-squared test of independence was used to test whether the proportion of individuals with neutralising antibodies for spike and anti-RBD was similar across four age groups. Neutralising antibody concentrations in each age group, for wild type, alpha, beta, and gamma variants of spike and anti-RBD, were further compared using Kruskal-Wallis test (comparison between > 2 groups) and Wilcoxon test (pairwise-comparison between groups) with the Benjamini and Hochberg multiple comparisons test adjustment. The Shapiro-Wilk test was used to determine if the neutralising antibody concentration data significantly deviated from a normal distribution. The ‘outcode’ data, a partial UK postcode constituting the postcode area and the postcode district, available with each participant sample, was used to generate geographic maps of % reactivity and participant numbers. Outcode boundary shapes, the total number of individuals that tested positive for anti-SARS-CoV-2 IgG, and the total number of tests performed were used to calculate outcode-level % reactivity rates. Statistical analyses were performed using the R statistical software (version 3.6.3) in RStudio (version 1.2.5033).

## Results

Across three testing rounds spanning June-July 2020, November-December 2020 and June-July 2021 (rounds 1-3 respectively), a total of 4844 residual sera/plasma specimens were assayed for SARS-CoV-2 Ig using a combination of commercial kits sourced from Euroimmun and Roche. Initial analyses were conducted using the Euroimmun anti-SARS-CoV-2 ELISA with the spike S1 domain as target antigen, whereas subsequent analyses employed the Roche Elecsys assay to examine sample reactivity towards the nucleocapsid protein. This ensured that seroprevalence estimates were confined to natural infection only and not confounded by vaccinated individuals with induced antibody responses against the spike protein.

Our choice to sample at various timepoints enabled temporal assessment of antibody responses at different phases of the pandemic. Round 1 studies were performed following the first national lockdown that was enforced in March 2020 (post-lockdown 1), whereas rounds 2 and 3 coincided with the pre- and post-lockdown phases of the second national lockdown imposed in December 2020 (pre- and post-lockdown 2, respectively). Of the 1108 samples assayed during round 1, a total of 43 showed detectable levels of anti-SARS-CoV-2 Ig equating to a seropositivity rate of 3.9% (95% CI 2.9%-5.4%) (**Supplementary Table 1 and Supplementary Table 2**). This increased only marginally during round 2, where 5.2% (95% CI 4.0%-6.5%) of samples were seropositive for anti-SARS-CoV-2 Ig (64 of 1242). However, a more pronounced increase was observed during round 3, where 11.6% (95% CI 10.4%-13.0%) of samples were seropositive for anti-SARS-CoV-2 Ig (290 of 2494).

Variable antibody responses were also observed in different demographic cohorts segregated according to age, gender, Health and Social Care Trust or place of residence (**Supplementary Table 3**). Collective sera for the entire study were sampled from a wide age range of 0 – 100 years, although the majority were from persons aged 50+ years with comparatively fewer from those <50 years during each testing round (**Supplementary Table 3**). Clear age-related differences in antibody responses were observed at various stages of the pandemic. For example, during round 1 of testing (post-lockdown 1), seropositivity rates were marginally lower in those aged 0 – 29 years at 2.7% compared to other age brackets (**Figure 1**). However, subsequent testing rounds revealed a shift in this trend as seropositivity rates became higher in the younger age brackets, peaking at 16.1% in 0 – 29 year olds during round 3 of testing (post-lockdown 2) (**Figure 1**).

**Figure 1.**
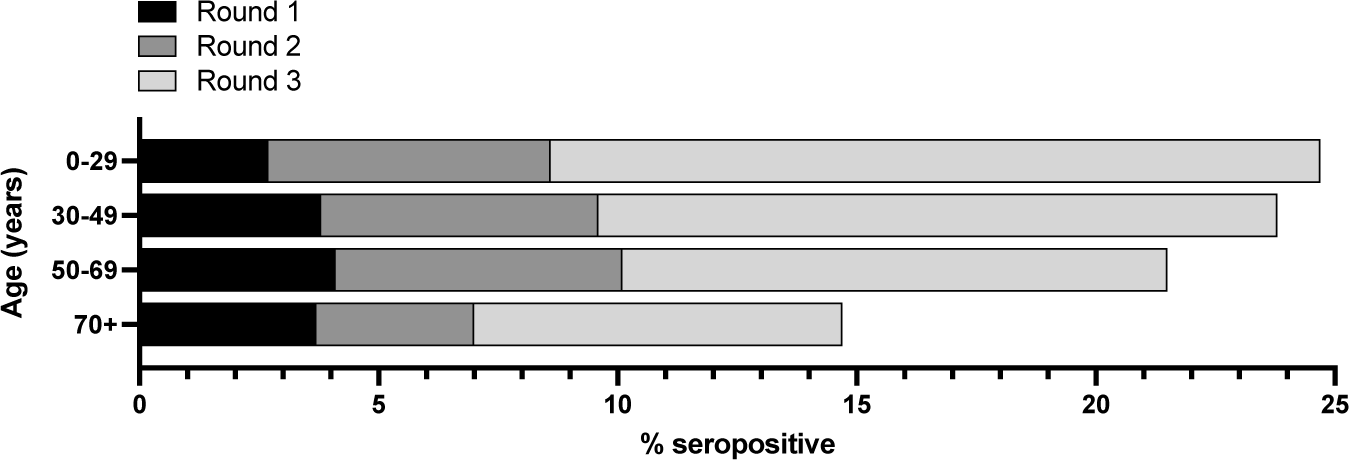
Age breakdown of SARS-CoV-2 Ig seroprevalence for each testing round. Data presented as % of sera within each age bracket that tested positive for anti-SARS-CoV-2 Ig.

The anti-IgG and anti-RBD IgG neutralising concentrations in round 3 sera/plasma showed age dependent increases for spike proteins (**Figure 2**). The majority of participants determined to be S-antibody reactive (by anti-SARS-CoV-2 IgG assay) had neutralising antibodies above the reactive cut-off for wild type (WT) spike (70.1%) and S1 RBD (73.4%) (**Supplementary Table 4**). Amongst the variants tested, higher proportions of the S-antibody reactive population showed neutralising antibody reactivity toward the alpha versions of S1 RBD (72.7%) and spike (64.4%) proteins. In addition, significant differences in the proportion of S-antibody reactive individuals with neutralising antibodies for WT (p < 0.001), and alpha (p < 0.001), beta (p = 0.001) and gamma (p < 0.001) variants of spike protein were found between age groups. We observed no significant age differences in the proportion of subjects with neutralising antibodies targeting WT (p = 0.22), and alpha (p = 0.26), beta (p = 0.06), and gamma (p = 0.08) variants of S1 RBD. However, significant differences in neutralising antibody concentrations between age groups were observed both for the wild type (p < 0.001) and the alpha (p = 0.008) variant of spike protein and the wild type (0.044) and the alpha (0.037) variant of S1 RBD. The pairwise-comparison revealed statistically significant differences between: i) 0-29 and 50-69 age groups (p = 0.033), 0-29 and 70+ age groups (p = 0.033), 30-49 and 50-69 age groups (p = 0.004), and 30-49 and 70+ age groups (p = 0.004) for the wild type of spike protein; ii) 0-29 and 50-69 age groups (p = 0.047), 0-29 and 70+ age groups (p = 0.022), and 30-49 and 70+ age groups (p = 0.045) for the alpha variant of spike protein; iii) 0-29 and 70+ age groups (p = 0.041) and 30-49 and 70+ age groups (p = 0.041) for the wild type of S1 RBD; and iv) 30-49 and 70+ age groups (p = 0.043) for the alpha variant of S1 RBD.

**Figure 2.**
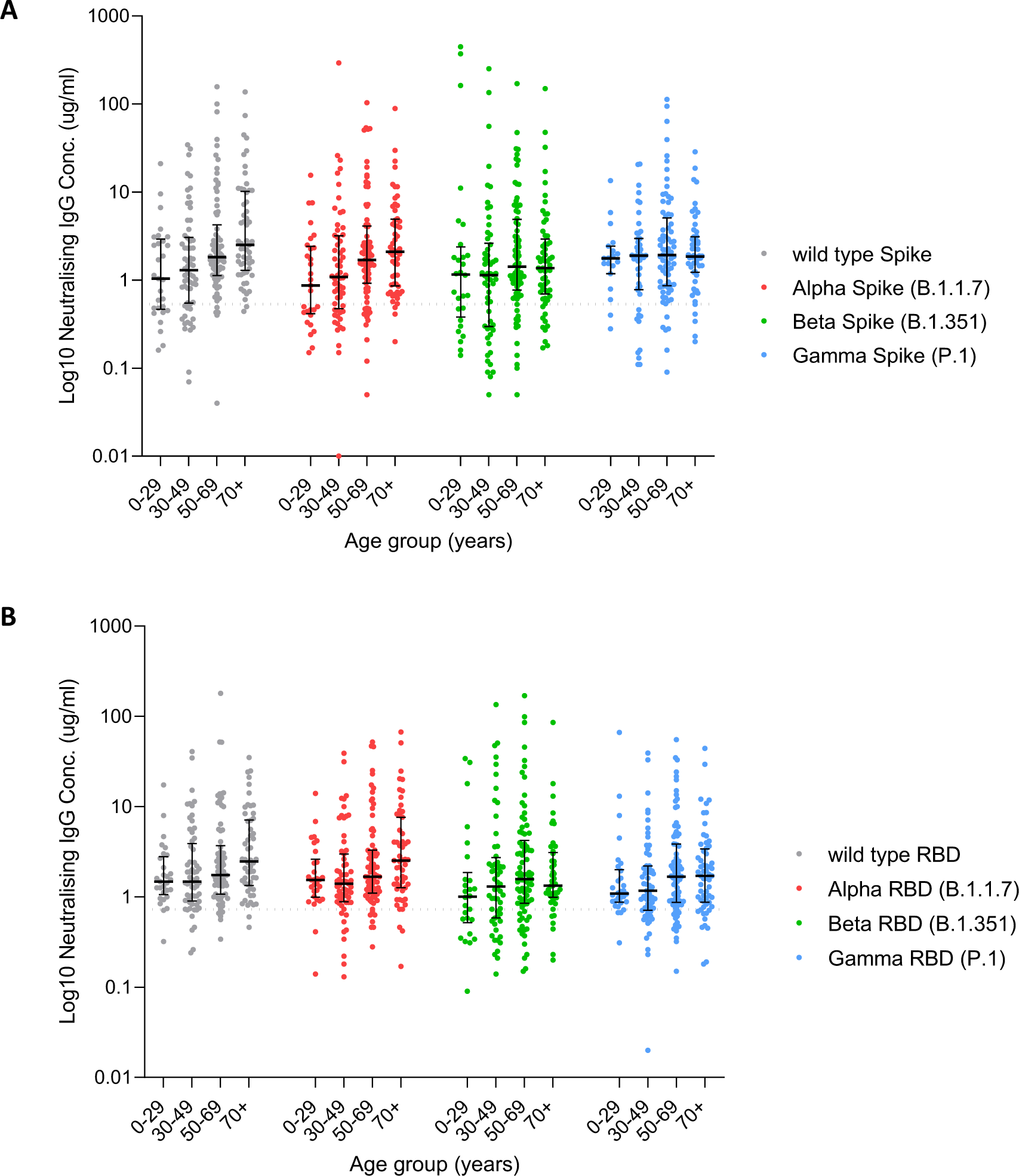
Neutralising IgG concentration of round 3 subpopulation. Neutralising antibody concentration (μg/ml) for a subpopulation of n=278 round 3 samples split by age group for wild type (grey dots), alpha (red), beta (green) and gamma (blue) variants of (A) spike protein and (B) receptor binding domain (RBD) of the S1 portion of the spike protein. Central horizontal bar is median, error bars are inter-quartile range; dotted line indicates reactivity cut off concentrations for (A) spike = 0.533

Variations in antibody response were also observed between genders as the pandemic progressed, with males showing higher seropositivity rates than females during rounds 1 (4.2% *vs.* 3.4%) and 2 (5.4% *vs.* 5.0%) of testing, whereas this trend was reversed in round 3 (9.9% *vs.* 12.9%) (**Figure 3**). Of note, however, a much larger number of females were sampled versus males during all three testing rounds (**Supplementary Table 3**).

**Figure 3.**
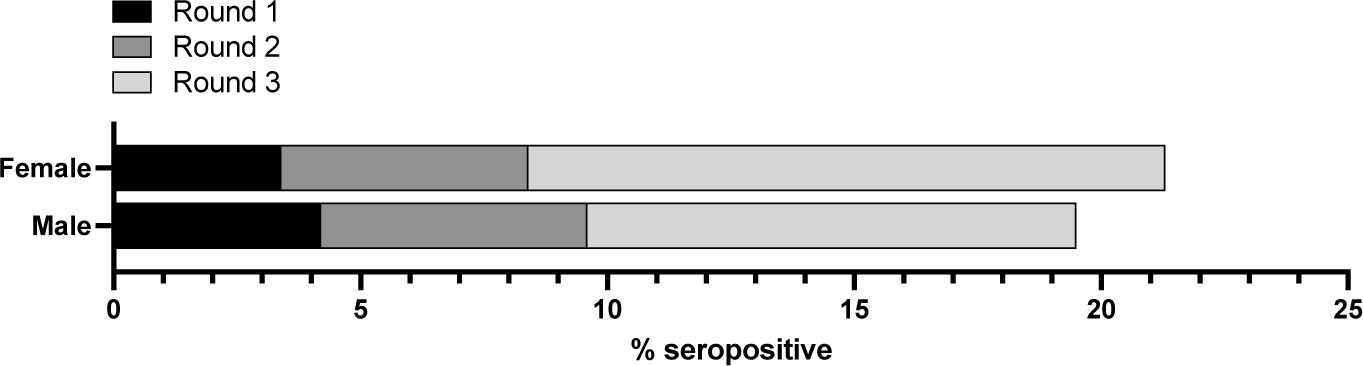
Gender breakdown of SARS-CoV-2 Ig seroprevalence for each testing round. Data presented as % of sera within each gender that tested positive for anti-SARS-CoV-2 Ig.

In terms of Health and Social Care Trust, a comparable number of samples were assayed from each of the Belfast, Northern, South Eastern, Southern and Western Trusts during all three testing rounds (**Supplementary Table 3**). All Trusts recorded the highest seropositivity rates during round 3 of testing (**Figure 4A**). For the Belfast, Northern and Western Trusts, seropositivity increased with each sequential testing round. However, the Southern and South Eastern Trusts showed an initial decline in seropositivity during round 2 versus round 1 of testing, which was reversed during round 3.

**Figure 4.**
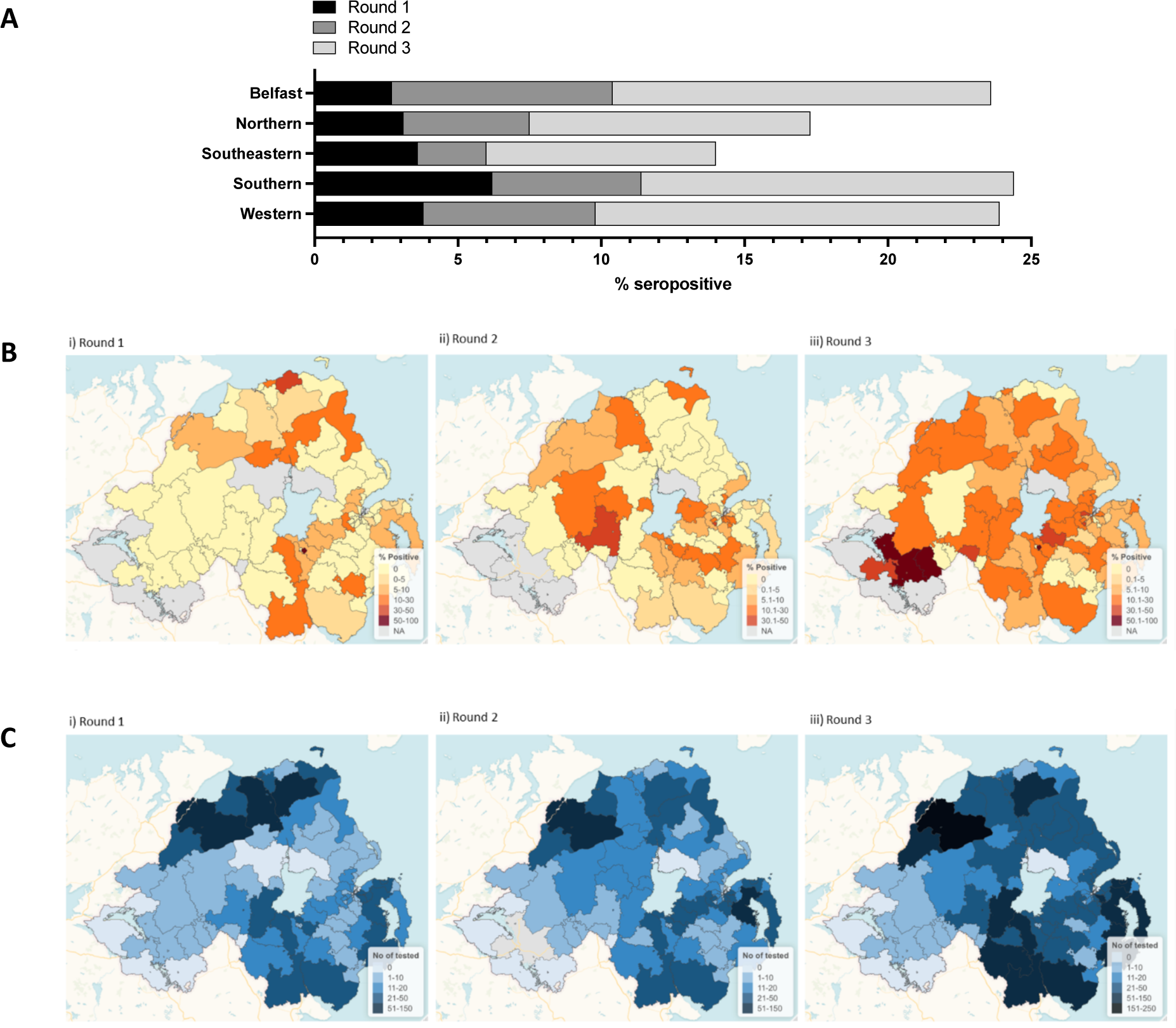
Geographical breakdown of SARS-CoV-2 Ig seroprevalence for each testing round. (A) Health and Social Care Trust breakdown of SARS-CoV-2 Ig seroprevalence for each testing round. Data presented as % of sera within each Trust that tested positive for anti-SARS-CoV-2 Ig. (B-C) Outcode maps of % reactivity and samples tested. Increased colour density indicates: (B) percentage of the population reactive by IgG immunoassay and (C) numbers of individuals tested, per outcode region for each sampling round (i-iii).

Further analysis of the outcode-level % reactivity rates, showed outcode-related differences in antibody responses over the three rounds of study (**Figure 4B-C**). For round 1 of testing, overall seropositivity rates were highest in BT65 (Legahory, Moyraverty, Tullygally, Drumgor, Monbrief), BT62 (Craigavon, Portadown, Tandragee, Clare, Scotch Street), and BT17 (West Belfast, North Lisburn) postcode areas at 100.0%, 22.2% and 15.4% respectively. However, the number of participants differ significantly between postcodes, with the number of samples assayed from BT65, BT62, and BT17 at 1, 27, and 13 respectively. During round 2, the highest seropositivity rate of 33.3% was recorded for BT10 (South Belfast) and BT70 (Dungannon, Ballygawley, Cappagh, Castlecaulfield, Donaghmore, Galbally, Garvaghy, Pomeroy, Rock, Seskilgreen) postcodes. Although the BT65 postal area showed an initial decline in seropositivity during round 2 versus round 1 of testing, round 3 revealed a shift in this trend, with BT65 having again one of the highest seropositivity rates (55.6%) in Northern Ireland. During round 3, the BT94 postcode area (Irvinestown, Ballinamallard, Brookeborough, Tempo, Maguiresbridge, Lisbellaw) reported the highest proportion of individuals that tested positive for anti-SARS-CoV-2 IgG. For the following postcodes, seropositivity increased with each sequential testing round: BT4 (East Belfast), BT7 (South Belfast), BT11 (West Belfast), BT24 (Ballynahinch, Drumaness, Saintfield), BT28 (Lisburn, Ballinderry Lower, Ballinderry Upper, Stoneyford), BT30 (Downpatrick, Ardglass, Ballyhornan, Ballykinler, Castleward, Clough, Crossgar, Kilclief, Killard, Killough, Killyleagh, Listooder, Loughinisland, Seaforde, Strangford, Toye), BT34 (Newry, Annalong, Ballymartin, Cabra, Hilltown, Kilcoo, Kilkeel, Mayobridge, Rathfriland, Rostrevor, Warrenpoint), BT37 (Newtownabbey), BT47 (Derry, Waterside, Claudy, Feeny, Dungiven, Eglinton, Park, New Buildings), BT49 (Limavady, Ballykelly), BT66 (Derryadd, Derrytrasna, Dollingstown, Donaghcloney, Lurgan, Waringstown), and BT82 (Strabane, Artigarvan, Ballymagorry, Bready, Clady, Douglas Bridge, Dunamanagh, Sion Mills, Victoria Bridge).

## Discussion

Following the emergence of SARS-CoV-2, it was rapidly acknowledged that surveillance programs would require urgent implementation to monitor virus activity and inform policy for many years ahead. In response, our study was initiated to gather information on the seroprevalence of anti-SARS-CoV-2 Ig in Northern Ireland, which would provide an indication of those with previous exposure to the virus. Residual sera from clinical biochemistry laboratories were assayed at various timepoints throughout 2020-2021 using a combination of anti-SARS-CoV-2 immunoassays against both the spike and nucleocapsid antigens. Our analysis showed that seroprevalence rates increased as the pandemic progressed and were also strongly influenced by demographics including age, gender and place of residence.

A limited number of studies have investigated SARS-CoV-2 seroprevalence in Northern Ireland to date. These include the ongoing COVID-19 Infection Survey, which was initially piloted in England and subsequently extended to other devolved UK nations including Northern Ireland in September 2020. A subset of the study participants provide blood samples for antibody testing, allowing seroprevalence rates to be closely tracked with updates published at regular intervals. Data indicated that in November 2020, an estimated 3.3% (95% CI: 1.6-6.0%) of the Northern Ireland population would have tested seropositive for anti-SARS-CoV-2 Ig, rising to 7.8% (95% CI: 3.5-14.4%) in December 2020.^16–17^ In comparison, the same time period was captured during round 2 of our study where seroprevalence was estimated at 5.2% (95% CI: 4.0-6.5%). These figures are surprisingly similar despite the notable difference in sourcing sera for both studies; either from hospital-based clinical biochemistry laboratories in our case, or from randomly selected private residential households throughout the general population for the COVID-19 Infection Survey. In another UK-wide multicentre study conducted between April to July 2020, a seroprevalence of 0.9% (95% CI: 0.2-3.3%) was reported in children (aged 2-15 years, N=215) of healthcare workers based in Belfast.^18^ This timeframe coincided with round 1 of our study, where 12.5% of samples within this age bracket tested positive for anti-SARS-CoV-2 Ig, although the low numbers involved (N=8) limit the robustness of these findings.

In the round 3 cohort, neutralising antibodies were most frequently detected to spike and S1 RBD of the alpha (B.1.1.7) variant across most age groups, which was the predominant strain in the preceding winter and spring months of 2021.^19^ Since no record of vaccination or prior SARS-CoV-2 infection was available, a more thorough interpretation of neutralising antibody persistence was not possible. However median concentration of neutralising antibodies directed toward spike protein WT and alpha variants were elevated in the 50-69 year olds relative to other age groups, whereas median concentrations of those directed against the S1 RBD WT and alpha variants were observed at higher levels in the over 70 year old group only. Also alpha variant specific neutralising antibodies were more frequently detected in the under 30 age group against S1 RBD than spike. These findings need confirmation in larger sized age groups with full vaccination and infection history, if high throughput ACE2 pseudoneutralisation assays are to be used to predict population protective immunity. The same ACE2 pseudoneutralisation assay has indicated that previous infection removed any negative influence of age or immune senescence on the level and quality of vaccine-induced immune responses in care home staff and patients.^20^ However the round 3 data provide initial insight into age related variations in neutralising antibody diversity in response to likely variant virus infection in the under 30s in particular, along with vaccination rollout prioritised to the over 50s, in the months preceding sample collection.^21^

Beyond Northern Ireland, there are abundant reports of SARS-CoV-2 seroprevalence studies, totalling 3924 in 140 countries as listed on the SeroTracker database^22^ as of December 12^th^ 2022 that have formed the basis of many comprehensive systematic reviews and meta-analyses.^23–24^ Significant diversity exists amongst these studies, with many focusing on defined study populations such as healthcare personnel,^25^ nursing home residents,^26^ schoolchildren,^27^ blood donors^28^ or those with comorbidities^29^ that limit the ability to draw meaningful comparisons with our own data. However, similar to our sampling methodology, several studies also make use of existing residual sera from healthcare settings.^30–31^ For example, a Scottish study between April and June 2020 reported a seroprevalence of 4.3% (95% CI: 4.2-4.5%) in residual samples from those accessing primary care via general practice.^32^ This timeframe approximately correlated with round 1 of our study, where a similar seroprevalence of 3.9% (95% CI: 2.9-5.4%) was observed.

Our choice of a ‘convenience’ sampling approach may have led to biased estimates of seroprevalence in our study. We identified residual sera from clinical biochemistry laboratories across Northern Ireland as a feasible sampling source, given that the routine processing of blood in this setting would facilitate access to sufficient sample numbers to address the urgent need for seroprevalence insights. However, with sera acquired from those accessing healthcare, it is possible that many of these individuals may have comorbidities that placed them at higher risk of severe infection. Such individuals may have been more likely to comply with preventive public health measures, potentially leading to a reduced incidence of infection as already noted in several publications.^33–34^ Nonetheless, we observed comparable trends to the ONS survey as discussed previously, where a more robust sampling strategy was employed.

A further limitation of our work relates to the demographic profile of the study population, which was not wholly representative of the Northern Ireland general population. Recent census data indicates that males and females comprise 49% and 51% of the Northern Ireland population, respectively. Gender misrepresentation was therefore apparent in our study, since sera were acquired from a disproportionate number of females (N=2679) versus males (N=2160). Moreover, the age demographic of our study cohort did not reflect the Northern Ireland population at large, with 66.6% of samples acquired from persons of 50+ years despite this age bracket only constituting 36.77% of the overall population.^35–36^ Although our estimates were adjusted in line with the true age and gender distribution of the Northern Ireland population, this may not fully mitigate sampling bias in our study. Our findings must therefore be interpreted with caution, especially in light of accumulating evidence that demographic characteristics can influence susceptibility to COVID-19.^38–40^

In conclusion, our findings highlight the evolving trajectory of SARS-CoV-2 in Northern Ireland, with seropositivity rates of <15% at three distinct timepoints suggesting that a significant proportion of the population had not yet naturally contracted the virus by mid-2021. Moreover, we report varying trends in seropositivity based on demographic factors, including age in particular where the highest rates were recorded in older generations in mid-2020 before a shift to younger generations in mid-2021. Despite sampling constraints, our study fulfilled the critical need for seroprevalence data following the outbreak of SARS-CoV-2, providing timely insights into virus transmission to inform planning and public health policy in Northern Ireland.

## Declarations

### Ethical approval and consent to participate

Ethical approval for this study was obtained from the Northern Ireland Biobank. The Northern Ireland Biobank has ethical approval from The Office of Research Ethics Committees Northern Ireland (ORECNI REF 21/NI/0019) and can confer ethical approval for projects which have received material from the bank.^14^

### Consent for publication

Not applicable

### Data availability

The datasets used and/or analysed during the current study are available from the corresponding author on reasonable request.

### Author Contribution

JSE, TF, AJB, JMcC, JB, DSG and CJS contributed to conception and design of the study. MKG, PS, AE, JMcL, MB, TSR, MMcD, AW, GB, ML, KD, GD, CL, JAJ and MOK contributed to acquisition of data. MKG, PS, AE, MB, JMcL, CC, TSR, AJB, DSG and CJS contributed to analysis and interpretation of data. MKG, PS, DSG, CC and CJS drafted the article. All listed authors provided final approval of the version to be submitted.

## Acknowledgements

Not applicable

