## Supplementary for "Analysis of SARS-CoV-2 Ig seroprevalence in Northern Ireland"

**Analysis of SARS-CoV-2 Ig seroprevalence in Northern Ireland. - Supplementary material**

***Supplementary Table 1. Overall estimates by time period.***

| | June/July 2020 | | Nov/Dec 2020 | | June/July 2021 | |
|---|---|---|---|---|---|---|
| | cases/total | estimate (95% CI/CrI) | cases/total | estimate (95% CI/CrI) | cases/total | estimate (95% CI/CrI) |
| Crude apparent proportion | 43/1108 | 3.9% (2.9%, 5.4%) | 64/1242 | 5.2% (4.0%, 6.5%) | 290/2494 | 11.6% (10.4%, 13.0%) |
| Standardised apparent proportion[1] | 43/1108 | 3.6% (1.7%, 5.6%) | 64/1238 | 5.2% (3.6%, 6.7%) | 290/2493 | 12.9% (11.2%, 14.5%) |
| Crude true proportion[2] | 43/1108 | 3.4% (1.6%, 5.2%) | 64/1242 | 5.1% (3.2%, 7.1%) | 290/2494 | 13.4% (11.4%, 15.7%) |
| Standardised true proportion[3] | 43/1108 | 3.6% (1.8%, 5.8%) | 64/1238 | 5.4% (3.7%, 7.5%) | 290/2493 | 14.5% (12.5%, 16.7%) |

[1]Based upon NI age (0-29,30-49,50-69,70+) and gender distribution, one individual missing gender. [2]True proportion using Bayesian method (used by PHE England), credible intervals used instead of confidence intervals, assuming sensitivity of 79% (137/173) and specificity of 98% (699/707). [3]True proportion using Bayesian method (used by PHE England), credible intervals used instead of confidence intervals, assuming sensitivity of 79% (137/173) and specificity of 98% (699/707) and standardised based upon NI age (0-29,20-49,50-69,70+) and gender distribution.

***Supplementary Table 2. Comparison of COVID-19 proportions by time period.***

| | COVID-19 cases/total (%) | P[1] | Odds Ratio (OR) (95% CI) | P | Adjusted[2] OR (95% CI) | P |
|---|---|---|---|---|---|---|
| June/July 2020 | 43/1108 (3.9%) | | 1.00 (ref. cat.) | | 1.00 (ref. cat.) | |
| Nov/Dec 2020 | 64/1242 (5.2%) | <0.001 | 1.35 (0.91, 2.00) | 0.14 | 1.33 (0.89, 1.99) | 0.16 |
| June/July 2021 | 290/2494 (11.6%) | | 3.26 (2.35, 4.53) | <0.001 | 3.09 (2.21, 4.31) | <0.001 |

[1]Chi-squared 2x3 P-value. [2]Adjusting for trust, age and sex.

***Supplementary Table 3. Sample characteristics by time period.*** Breakdown of total numbers of samples assayed (N) and percentage (%) according to Health and Social Care Trust, age group and gender across all three testing rounds.

| | Round 1<br>June/July 2020<br>N (%) | Round 2<br>Nov/Dec 2020<br>N (%) | Round 3<br>June/July 2021<br>N (%) |
|---|---|---|---|
| Health and Social Care Trust | | | |
| Belfast | 223 (20.1%) | 246 (19.8%) | 499 (20.0%) |
| Western | 210 (19.0%) | 248 (20.0%) | 495 (19.8%) |
| Southern | 225 (20.3%) | 248 (20.0%) | 500 (20.0%) |
| Northern | 225 (20.3%) | 250 (20.1%) | 500 (20.0%) |
| South Eastern | 225 (20.3%) | 250 (20.1%) | 500 (20.0%) |
| Age group | | | |
| 0-29 | 74 (6.7%) | 152 (12.3%) | 341 (13.7%) |
| 30-49 | 185 (16.7%) | 278 (22.4%) | 585 (23.5%) |
| 50-69 | 394 (35.6%) | 452 (36.5%) | 850 (34.1%) |
| 70+ | 453 (41.0%) | 357 (28.8%) | 718 (28.8%) |
| Gender | | | |
| Female | 580 (52.5%) | 682 (55.0%) | 1417 (56.8%) |
| Male | 525 (47.5%) | 559 (45.0%) | 1076 (43.2%) |

***Supplementary Table 4. Neutralising IgG reactivity summary of round 3 subpopulation.*** *Reactivity cut-off concentrations: * Spike =0.533 $\mu$g/ml; $ S1 RBD =0.726 $\mu$g/ml.*

| | | | **0-29 years** (n=35) | | **30-49 years** (n=80) | | **50-69 years** (n=99) | | **70+ years** (n=64) | | **Total** (n=278) | |
|---|---|---|---|---|---|---|---|---|---|---|---|---|
| | | Age (Median, IQR) | **22** | **± 3.75** | **40.5** | **± 10** | **59** | **± 8** | **78** | **± 8.5** | **55** | **± 29.25** |
| | | Female (n, %) | 17 | 48.6% | 47 | 58.8% | 54 | 54.5% | 38 | 59.4% | 156 | 56.1% |
| **ACE2 pseudoneutralisation assay** | **wild type** | Spike Reactive (n, %)* | 19 | 54.3% | 48 | 60.0% | 73 | 73.7% | 55 | 85.9% | 195 | 70.1% |
| | | S1 RBD Reactive (n, %)$ | 27 | 77.1% | 53 | 66.3% | 72 | 72.7% | 52 | 81.3% | 204 | 73.4% |
| | **S1 RBD variant$** | Alpha (B.1.1.7) Reactive (n, %) | 28 | 80.0% | 54 | 67.5% | 69 | 69.7% | 51 | 79.7% | 202 | 72.7% |
| | | Beta (B.1.351) Reactive (n, %) | 16 | 45.7% | 39 | 48.8% | 65 | 65.7% | 39 | 60.9% | 159 | 57.2% |
| | | Gamma (P.1) Reactive (n, %) | 26 | 74.3% | 47 | 58.8% | 72 | 72.7% | 49 | 76.6% | 194 | 69.8% |
| | **Spike variant*** | Alpha (B.1.1.7) Reactive (n, %) | 16 | 45.7% | 44 | 55.0% | 66 | 66.7% | 53 | 82.8% | 179 | 64.4% |
| | | Beta (B.1.351) Reactive (n, %) | 19 | 54.3% | 40 | 50.0% | 67 | 67.7% | 51 | 79.7% | 177 | 63.7% |
| | | Gamma (P.1) Reactive (n, %) | 17 | 48.6% | 38 | 47.5% | 70 | 70.7% | 50 | 78.1% | 175 | 62.9% |
| **Roche Elecsys Anti-SARS-CoV-2 IgG Assay** | | N-Antibody Reactive (n, %) | 2 | 5.7% | 5 | 6.3% | 7 | 7.1% | 1 | 1.6% | 15 | 5.4% |
| | | S-Antibody Reactive (n, %) | 16 | 45.7% | 53 | 66.3% | 92 | 92.9% | 58 | 90.6% | 219 | 78.8% |
